# Comparing Three Approaches to Modelling the Effects of Temperature and Rainfall on Malaria Incidence in Different Climatic Regions

**DOI:** 10.1101/2024.07.19.24310710

**Authors:** Gladstone T. Madito, Basil D. Brooke, Sheetal P. Silal

## Abstract

**Backgroun:** Malaria transmission is primarily limited to tropical regions where environmental conditions are conducive for the survival of *Plasmodium* parasites and *Anopheles* mosquitoes. Adequate rainfall provides breeding sites, while suitable temperatures facilitate vector mosquito life-cycles and parasite development. Evaluating the efficacy of vector control interventions is crucial to determine their effectiveness in reducing malaria transmission. The aim of this study was to explore how these factors affect transmission dynamics at varying levels of vector control efficacy.

**Methods:** We developed a vector-host compartmental mathematical model to compare three published approaches to incorporating weather influences on malaria transmission. The first approach examines mosquito biting behaviour and mortality rates in larval and adult stages. The second focuses on temperature effects on mosquito life-cycle characteristics throughout the aquatic and adult stages. The third considers how temperature and rainfall influence adult mosquito behaviour, environmental carrying capacity, and survival during the aquatic stages. Model simulations were conducted at different annual vector control coverage levels, to identify variations in transmission patterns and seasonal variability in daily and annual incidence across three climate regions.

**Results:** The first approach indicates sustained seasonal transmission, with lower cases per 1,000 in tropical regions compared to semi-arid and sub-tropical regions, even with enhanced vector control reducing cases. The second approach projects extended seasonal peaks in malaria transmission in tropical and semiarid regions, driven by prolonged warm periods, while sub-tropical regions show lower incidence due to cooler temperatures limiting mosquito survival. In contrast, the third approach projects multiple irregular peaks, with transmission ceasing in winter across all regions.

**Conclusions:** Simulations indicate that climatic events like heatwaves or flooding, can trigger mosquito population surges and malaria outbreaks, even in areas previously free of malaria, despite strong vector control efforts. However, the results demonstrate that sustained and effective vector control, particularly in regions with moderate temperatures, can substantially reduce malaria incidence. Effective malaria control requires incorporating weather predictions into intervention plans, and enhancing current vector control strategies with supplementary measures like larval source management. Accurate timing and targeting of these interventions, based on transmission season projections, are crucial for maintaining robust control as weather conditions evolve and to prepare for future challenges.

## Background

Malaria is an infectious disease caused by *Plasmodium* parasites transmitted to humans through the bites of infectious female *Anopheles* mosquitoes [1]. This disease poses a significant public health concern, impacting the health and livelihoods of individuals in tropical regions worldwide [2–4]. Malaria remains a significant cause of mortality, with around half a million deaths attributed to the disease annually [5–7]. The World Health Organization (WHO) categorises the prevalence of malaria in a country based on the number of reported cases per population at risk. Countries with fewer than 100 cases per 1000 population per year are classified as low transmission, while those reporting more than 450 cases per 1000 population per year are considered high transmission settings [8]. Several intervention strategies have been implemented to combat malaria transmission, these include case management through effective treatment with artemisinin-based combination therapies (ACTs) together with vector control measures such as indoor residual spraying (IRS) of insecticides and widespread distribution of long-lasting insecticide-treated nets (LLINs) [9–16]. These interventions play a crucial role in reducing the incidence of malaria and improving public health outcomes in affected areas. Despite this, seasonal outbreaks frequently occur due to favourable weather conditions and reduced efficacy of interventions. The influence of temperature and rainfall on factors such as mosquito abundance or changes in mosquito breeding, survival and behaviour contribute to these challenges [2, 3, 17–19].

Malaria transmission is primarily restricted to tropical regions where temperatures are conducive for both parasite and mosquito development, and adequate rainfall to facilitate the availability of breeding sites for mosquitoes [9–11]. Mosquito breeding involves the laying of eggs in different water bodies, from temporary puddles to permanent lakes and dams with varying levels of salinity, turbidity and pollution. After hatching, the larvae undergo four developmental stages, known as instars, before they pupate. These life stages are aquatic, while adult mosquitoes are the only terrestrial phase, having emerged from the pupae. The rate of maturation from one stage to the next is primarily dependent on temperature [20]. Higher temperatures can accelerate mosquito development within their upper thermal limits, leading to a higher likelihood of survival during the aquatic stages and an increase in the adult mosquito population. Similarly, the malaria parasite matures more quickly within mosquitoes at warmer temperatures, with optimal sporogony occurring around 25_0_C [21]. However, if temperatures exceed the upper thermal limit of 30_0_C, sporogony is halted, and neither the mosquitoes nor the malaria parasite are likely to survive [2, 3, 6, 7, 21, 22]. Additionally, adequate rainfall generates sufficient habitats for mosquito larvae to grow, leading to abundant adult mosquitoes capable of spreading the disease. However, limited rainfall results in fewer breeding sites and heavy rainfall leads to flushing out of mosquito larvae, both of which result in reduced mosquito populations available to spread the disease [12, 14–16]. It is important to consider the impact of weather factors like temperature and rainfall on mosquito development, survival, and behaviour when implementing vector control measures to manage malaria outbreaks. Accurate seasonal weather forecasts of these factors can be used in malaria models as early warning systems in endemic regions. These models can also assess potential changes in malaria prevalence due to seasonal weather variations.

Several mathematical models incorporating empirical data and statistical approaches have shown associations between variations in meteorological factors with malaria incidence [5, 13, 17, 19, 23–25]. While extensive research into the influence of temperature variability on malaria transmission has been conducted, rainfall has often been relegated to a secondary factor, despite efforts to develop suitable models that account for the combined impact of temperature and rainfall on vector and parasite development [2, 3, 6, 26, 27]. The significance of these models has grown because while statistical models have been valuable in revealing relationships between environmental variables and transmission intensity, process-based mathematical models offer a more explanatory insight into the balance between internal factors (resulting from biological processes) and external factors (such as changes in environmental variables) that drive transmission. These dynamic models are essential because they account for the biological processes driving malaria transmission within an environment that changes over different time scales [24, 28–31]. Furthermore, a dynamic model is crucial for capturing invasion dynamics and effectively forecasting the emergence of new outbreaks. This includes scenarios where changes in temperature and rainfall patterns may render previously unsuitable areas conducive to transmission, as well as instances of human migration (and thereby *Plasmodium* parasite migration) or mosquito spread into previously unaffected regions [32–35]. A study in Mpumalanga province, South Africa, suggests that the adults of the malaria vector species *A. arabiensis* and *A. parensis* do not undergo a period of suspended development during the colder and drier winter months [36]. Instead, adult female mosquitoes continue to seek for blood meals in winter but their population densities are significantly reduced by the limited number of breeding sites available. Otherwise, colder temperatures likely reduce their blood feeding propensity and therefore their fertility, and as a result contribute to reducing transmission.

A mechanistic model calibrated with weather data from multiple regions in Africa found that malaria infection tends to rise within temperatures ranging from 16 to 25_0_C, and decreases within the 25 to 28_0_C range [31]. In contrast, a study using a climate-based vector-host model, which considers the aquatic stages in the mosquito lifecycle, indicated that transmission is optimized in the temperature range of 21 to 25_0_C, accompanied by 95 to 125 mm of rainfall [27]. A vector-host modelling approach was employed to investigate the dynamics of malaria transmission, specifically focusing on the aquatic stages of mosquito vectors [12]. The model categorized adult mosquitoes based on their behaviours such as biting, resting, and host-seeking, while also considering the impact of ambient air temperature and water body temperature on mosquito breeding success. By quantifying the seasonality of *Anopheles arabiensis* population densities, the model accurately simulated the observed trends in larval density [1–3, 12]. This alignment with laboratory experiments highlight the robustness of the model in capturing real-world conditions. In contrast, models that neglect the aquatic stages of mosquito vectors often fail to account for these crucial aspects of malaria transmission dynamics [5, 26, 35, 37].

While mechanistic models provide a valuable framework for understanding malaria dynamics, various approaches have been developed to incorporate the effects of temperature and rainfall on malaria transmission dynamics, focusing on different critical stages of transmission. [26] developed a weather-based model that examines malaria transmission by focusing on an age-structured vector population with periodic birth rates. Their model integrates factors such as temperature-dependent egg production, biting rates and mortality rates of both aquatic and adult mosquitoes. Similarly, [12] employed a weather-based mathematical model to investigate the impact of temperature on mosquito development and behaviours. This model incorporates temperature-dependent factors such as egg production, as well as the development and mortality rates of eggs, larvae and pupae, alongside adult mosquito activities such as resting, mating and host-seeking behaviours. Furthermore, [2, 22] utilized a weather-based mathematical model to explore the combined effects of temperature and rainfall on malaria transmission dynamics. Their model includes mosquito recruitment dependent on temperature and rainfall, infection dynamics between humans and mosquitoes, and explores the impact of interventions such as insecticide spraying. While previous studies have explored the impact of temperature and rainfall on malaria transmission, they typically address these factors in isolation or only consider specific stages of the mosquito life cycle. [26] do not explicitly consider aquatic stages, instead they consider the combined aquatic population as a single unit and assume that its mortality rate is equivalent to that of larvae. Additionally, [12] explicitly account for these stages but focus solely on mosquito population dynamics, neglecting transmission. Moreover, [2, 22] include the effects of rainfall but also neglect the aquatic population. Because these studies each focus on different aspects of malaria dynamics, it is essential to identify the most appropriate approach for investigating mosquito population dynamics and guiding resource allocation and decision-making for malaria control and prevention under varying weather conditions. This study aims to develop a compartmental mathematical model to evaluate the efficacy of vector control interventions by exploring how different approaches to incorporating weather influences affect malaria transmission dynamics. The research compares these approaches to identify the most effective framework for modelling the impacts of weather on malaria transmission and mosquito population dynamics across diverse settings. The study seeks to inform evidence-based strategies for optimizing vector control interventions, ensuring their timely and targeted implementation to sustain control and address emerging malaria transmission challenges under changing climatic conditions.

## Methods

We constructed a compartmental model by grouping individuals with similar characteristics related to developmental stage, risk of infection, infectiousness, treatment seeking behaviour and recovery from disease into compartments within mosquito and human populations. The model diagram (see Figure 1) illustrates the transmission of malaria between mosquito vectors and human hosts, accounting for the aquatic stages of the mosquito life-cycle which includes eggs (*E_a_*) that hatch into larva (*L_a_*) that develop into pupae (*P_a_*) that emerge as adult mosquitoes. We consider a collective population of larvae without explicitly accounting for the instar stages of larval development. The population of adult mosquitoes is grouped into mosquitoes susceptible to malaria infection (*S_m_*), infected mosquitoes that cannot transmit infection to humans (*E_m_*), and infectious mosquitoes that can transmit the infection to humans (*I_m_*). Those mosquitoes that cannot transmit infections can become infectious if *Plasmodium* sporozoites (following sporogony in the midgut oocyts) have migrated to their salivary glands. The host population is grouped into individuals at risk of infection (*S*), infected humans that cannot transmit the infection to mosquitoes (*E*), and infected people that can transmit the infection to mosquitoes but either do not show symptoms (*A*), show uncomplicated symptoms (*I_u_*) or are experiencing severe symptoms (*I_s_*). Furthermore, individuals treated for uncomplicated symptoms (*T_u_*) or severe symptoms (*T_s_*) and individuals that have recovered from the disease (*R*) are also considered.

**Fig. 1:**
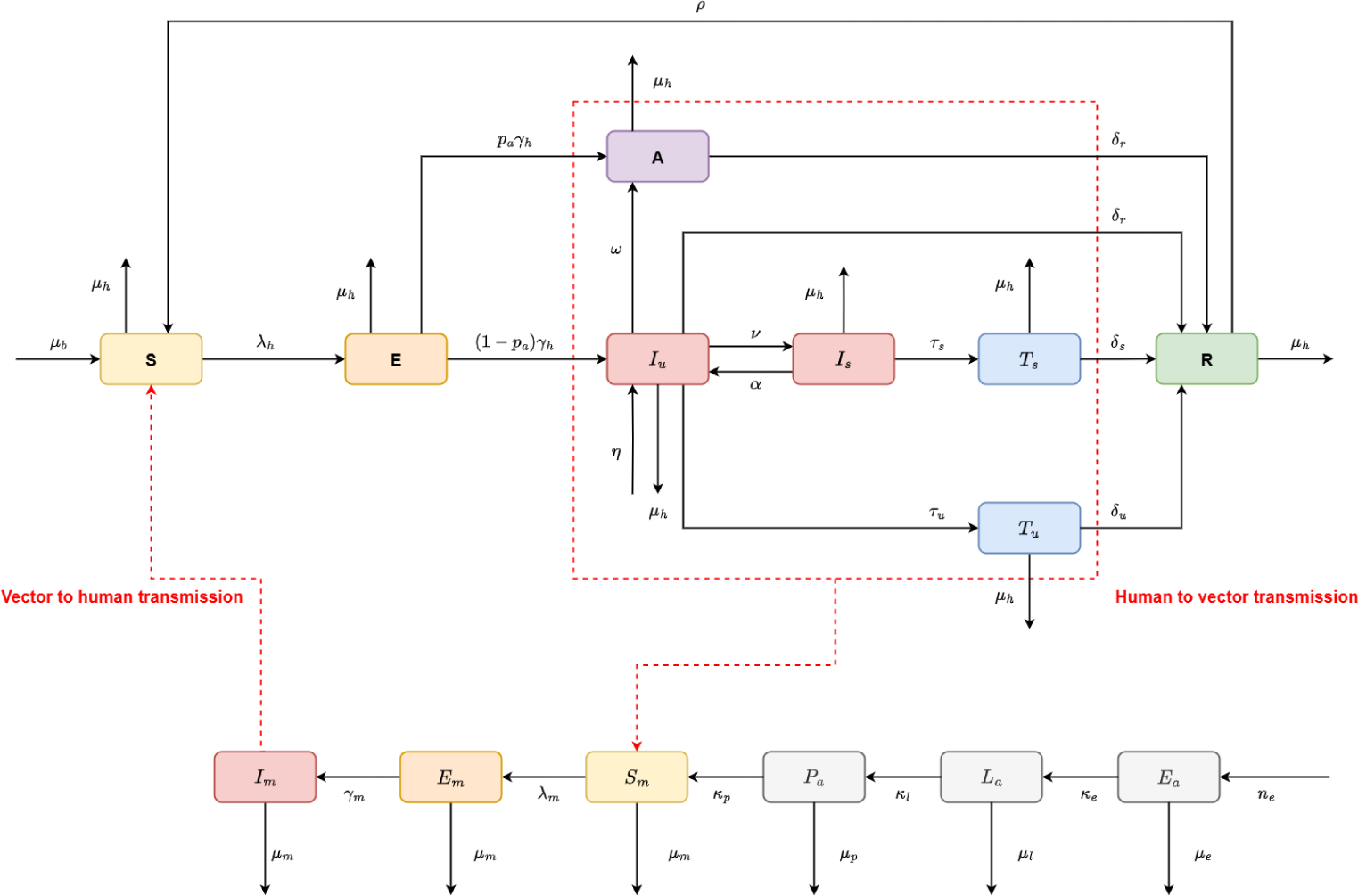
Transmission flow diagram illustrating the development of mosquitoes from egg to larvae then pupae and the transmission of malaria between a human and mosquito population.

We adopt the technique thoroughly discussed in [38], to investigate the role of vector control in effectively preventing transmission between mosquitoes and humans. In this modelling framework, vector control coverage is incorporated as a dynamic compartment that accounts for the annual deployment of interventions such as insecticide-treated nets (ITNs) and indoor residual spraying (IRS), as well as the gradual loss of their operational efficacy (see Appendix I for further details). The mosquito development and disease pathways illustrated in the model diagram in Figure 1 are represented by the system of ordinary differential equations (ODEs) describing the transmission of malaria between vector mosquitoes and human hosts (see Appendix I ODEs). Descriptions of the parameters and compartments are provided in Table 1 and Table 2.

**Table 1:** Vector and host compartments of the model (see Appendix I for mathematical equations).

| State | Description |
| --- | --- |
| $E_a$ | Mosquito eggs |
| $L_a$ | Mosquito larvae |
| $P_a$ | Mosquito pupae |
| $S_m$ | Mosquitoes susceptible to malaria infection |
| $E_m$ | Infected mosquitoes without sporozoites (exposed) |
| $I_m$ | Infectious mosquitoes |
| $S$ | Humans at risk of malaria infection |
| $E$ | Infected humans without gametocytes (exposed) |
| $A$ | Infectious humans without symptoms (asymptomatics) |
| $I_u$ | Infectious humans experiencing uncomplicated symptoms |
| $I_s$ | Infectious humans with severe malaria symptoms |
| $T_u$ | Patients treated for uncomplicated symptoms |
| $T_s$ | Patients treated for severe symptoms |
| $R$ | Recovered patients with temporary immunity acquired from infection |

**Table 2:** Descriptions of parameters. See Appendix I for values used to simulate the malaria transmission model.

| Symbol | Description |
| --- | --- |
| $n_e$ | Average number of eggs laid |
| $\kappa_e$ | Egg hatching rate |
| $\mu_e$ | Egg mortality rate |
| $\kappa_l$ | Larval development rate |
| $\mu_l$ | Larval mortality rate |
| $\kappa_p$ | Pupal development rate |
| $\mu_p$ | Pupal mortality rate |
| $\mu_m$ | Mortality rate in adult mosquitoes |
| $\gamma_m$ | Mosquito incubation rate |
| $\mu_b$ | Birth rate of humans |
| $\mu_h$ | Mortality rate in humans |
| $\delta_u$ | Recovery rate for uncomplicated symptoms |
| $\delta_s$ | Recovery rate for severe symptoms |
| $\gamma_h$ | Incubation rate within humans |
| $\nu$ | Development rate of severe symptoms |
| $\tau_u$ | Treatment rate for uncomplicated symptoms |
| $\omega$ | Recovery rate from severe symptoms |
| $\tau_s$ | Treatment rate for severe symptoms |
| $\mu_s$ | Disease-induced mortality rate |
| $\rho$ | Loss of temporary immunity |

### Model Compartments and Parameters

The transmission model is parameterized with values provided and fully described in Table 1 in Appendix I. The total number of mosquito eggs laid by a population of female mosquitoes is represented by *n_e_*, with these eggs being oviposited into water bodies by individual female mosquitoes at a rate of *θ* and cleared at a rate of *µ_e_*. Mosquito eggs hatch into larvae at a rate of *κ_e_*, and competition for nutrients and predation leads to larvae dying at a rate of *µ_l_*, whereas those that survive develop into pupae at a rate of *κ_l_*. Clearance of pupae from water bodies occurs at a rate of *µ_p_*and those that survives emerge as adult mosquitoes at a rate of *κ_p_*. Adult mosquitoes die at a rate of *µ_m_* and susceptible mosquitoes become infected at a rate of *λ_m_*. The malaria parasite develops from gametocytes to sporozoites within infected mosquitoes at a rate of *γ_m_*.

We consider that recruitment of individuals into the host population occurs at a birth rate of *µ_b_*, with the mortality rates in humans given by *µ_h_*. Susceptible humans become infected through infectious mosquito bites at a rate of *λ_h_* and malaria parasite development within individuals exposed to infection occurs at a rate of *γ_h_*. The proportion of infectious individuals that do not show symptoms is given by *pa* and the loss of uncomplicated symptoms occurs at a rate of *ω*. Infectious people with uncomplicated symptoms receive treatment at a rate of *τ_u_*, whereas those that do not receive treatment recover from disease at a rate of *δ_r_*. Infected people experience severe malaria symptoms at a rate of *ν* and are lost at a rate of *α*. People with severe symptoms receive treatment at a rate of *τ_s_* but suffer malaria induced death at rate of *µ_s_*. People that have been treated for uncomplicated or severe symptoms recover from disease at rates *δ_u_*and *δ_s_*, respectively. Finally, recovered people are considered to possess temporary immunity acquired from infection and lose this immunity to become susceptible to infection again at a rate of *ρ*.

### Comparison of Modelling Approaches

Three modelling approaches were adopted to assess the effects of temperature and rainfall on malaria transmission dynamics, each capturing different aspects of the mosquito life cycle and transmission process. The weather dependent functions were interpolated from laboratory-derived life history data for various *Anopheles* species, including *A. arabiensis, A. funestus* and others vectors such as *A. pseudopunctipennis, Culex quinquefasciatus, Aedes aegypti*, and *A. gambiae* complex. These functions describe mosquito development rates and behavioural responses across a range of temperatures [1, 2, 12, 22, 25, 39–45]. Although mosquito reproduction and maturation involve discrete biological events, the model uses continuous functions to represent the smooth, temperature-driven variation in key traits such as blood feeding propensity and fecundity. This allows a smoother integration into the model dynamics capturing population-level patterns rather than individual-level behaviour. Approach A focuses on aspects of transmission that are critical to the effectiveness of vector control that include mosquito biting behaviour, as well as mortality of adult mosquitoes and larvae. Vector control through IRS or ITNs limits contact between susceptible individuals and mosquitoes, which protects against infectious mosquito bites. Temperature also limit exposure to infected mosquitoes and the number of mosquitoes available to transmit infection by influencing the number of larvae that die and the lifespan of adult mosquitoes. These factors are also considered in Approach B which extends the role of temperature to the development and mortality of mosquito eggs, larvae, and pupae. The effects of temperature on the total number of eggs laid and the rate of ovipositing by a female mosquito during her lifespan are also considered. Despite overlap with both these approaches, Approach C accounts for the developmental period during aquatic stages and the survival probability of eggs, larvae and pupae in habitats with changing temperatures. Furthermore, temperature dependence of the extrinsic incubation period for parasites to develop within mosquitoes and rainfall dependence of the environmental carrying capacity are considered.

#### Approach A: Temperature-Regulated Mosquito Mortality

We adopt this approach to model three aspects of the mosquito life-cycle that are crucial to mosquito survival and malaria transmission. How many eggs survive the larval stage of the mosquito life-cycle to become pupae influences the growth of the adult mosquito population which is regulated through fluctuating mortality at different temperatures. Mosquitoes that survive into adulthood and come into contact with humans drive transmission through biting and blood feeding. Seasonal changes in temperature will affect this behaviour, especially when outside the optimal thermal limits of the mosquito population. It is assumed that mosquito larvae and adult mortality rates are at least partially temperature dependent, that is *µ_l_*(*T*) and *µ_m_*(*T*) for larvae and adult mosquitoes, respectively, and that the biting rate, *a*(*T*) (see Figure 9 in Appendix II for plots of these functions).

#### Approach B: Temperature-Sensitive Mosquito Life-Cycle

This approach extends our focus to account for the temperature dependence of mosquito characteristics related to the development of eggs, larvae and pupae are explicitly considered. We consider the total number of eggs laid by a population of female mosquitoes *n_e_*(*T*), the daily rate of egg oviposition *θ*(*T*), the development rates of eggs *κ_e_*(*T*), larvae *κ_l_*(*T*) and pupae *κ_p_*(*T*), the mortality rates of eggs *µ_e_*(*T*), larvae *µ_l_*(*T*), as well as the biting rate of adult mosquitoes *a*(*T*) (see Figure 10 in Appendix II for plots of these functions).

#### Approach C: Temperature and Rainfall Dependent Aquatic Survival

This last approach considers how the environmental carrying capacity of adult mosquitoes *K_m_*(*R*) and the survival probabilities of eggs *p_e_*(*R*), larvae *p_l_*(*R, T*) and pupae *p_p_*(*R*) depend on rainfall. The rainfall threshold for flushing out of aquatic mosquitoes due to heavy rainfall is denoted by *R_l_*. Furthermore, the biting rate *a*(*T*), mortality rate of aquatic and adult mosquitoes, *µ_l_*(*T*) and *µ_m_*(*T*) are included. The development period of larvae *t_l_*(*T*), and the incubation period of malaria parasites within mosquitoes *γ_m_*(*T*) are all considered temperature dependent (see Figure 11 in Appendix II for plots of these functions).

### Climate Data

This study uses historical mean monthly surface air temperature and precipitation datasets for the 1950 to 2022 period in three regions with semi-arid, tropical and sub-tropical climate conditions. Temperatures in the semi-arid region that had the highest peaks around 30 degrees Celsius, followed by tropical then the sub-tropical region which also experienced the lowest temperatures around 15 degrees Celsius during the 2016 to 2022 period. The tropical region accumulated the largest amount of rainfall peaking around 250 to 300 mm during months of heavy rainfall, followed by the semi-arid region with slightly less rainfall peaking around 150 to 250 mm despite experiencing the longest periods without rainfall. The sub-tropical region had the least amount of rainfall peaking below 150 mm during periods of heavy rainfall between 2016 and 2022. The temperature and rainfall trends for the period investigated in the study are presented in Figure 7 and 8 in Appendix II. The model is simulated with the same initial conditions and varying parameters across three modelling approaches A, B and C. Furthermore, we explore vector control scenarios with annual coverage levels ranging from 25 % to 90 % in three climate regions. Simulations are conducted for the entire period of seventy two years from 1950 to 2022 to observe annual trends in malaria incidence and variations in seasonal patterns are analysed for the last six years from 2016 to 2022. The impact of temperature and rainfall on malaria transmission in three regions are compared through projected aquatic and adult mosquitoes populations as well as the number of new cases, populations at-risk of infection, asymptomatic populations and annual incidence rates. We conducted our comparative study using formulations of the temperature and rainfall dependent model parameters derived in studies by [1, 26] for Approach A, [2, 12] for Approach C, and we performed a nonlinear least squares fitting process to validate the functions derived from *A. arabiensis* mosquito life traits data in [22] (see Appendix I for detailed explanation of the fitting process).

## Results

### Aquatic Mosquito Population Dynamics

Three approaches were adopted to integrate the influence of weather conditions on the development and survival of Anopheles mosquito eggs, larvae and pupae provided vector control intervention in these regions. Figure 2 shows that Approach A projects relatively lower aquatic mosquito populations throughout the year compared to Approaches B and C, with egg populations peaking around 12 million, whereas larvae peaked around 10 million and pupal populations remained below 3 million in all three regions. In addition, Approach A predicts persistent and stable aquatic mosquito populations, characterized by regular monthly population surges across all regions. Approach B, in contrast, projects rapidly fluctuating yet stable aquatic populations, with irregular multi-peaked seasonal growth patterns observed despite very low pupal abundance. Similarly, Approach C predicts a single annual surge in egg populations, occurring even in the presence of consistently low larval and pupal populations.

**Fig. 2:**
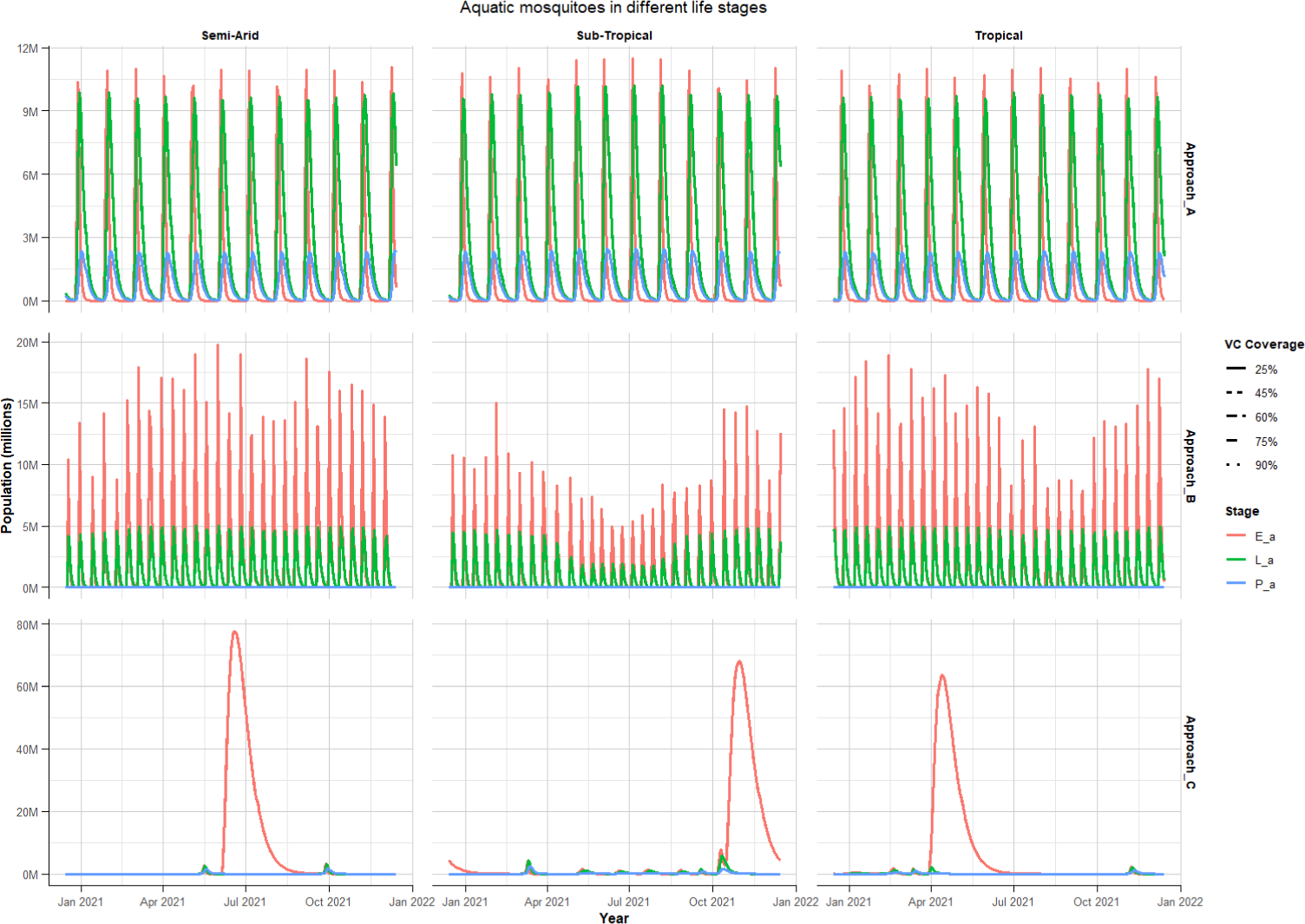
Projected aquatic mosquito populations across regions during the Jan 2021 to Jan 2022 period.

Approach B shows egg populations peaking below 20 million across all regions, despite differences in transmission dynamics. In the sub-tropical region, aquatic populations decline to around 5 million during the cooler June-July winter period, while egg populations show distinct seasonal dynamics with peaks of approximately 15 million. Although adult mosquito mortality is increased in warmer climates, higher temperatures accelerate oviposition and increase fecundity, thereby sustaining egg population growth in the semi-arid and tropical regions. Approach C on the other hand, indicates that adequate rainfall creates favourable aquatic habitats, enabling mosquito eggs, larvae, and pupae to persist across a wide range of temperatures throughout the year. This approach projects distinct seasonal peaks in egg populations, reaching approximately 60 million in the tropical region, 75 million in the sub-tropical region, and 80 million in the semi-arid region. Furthermore, Approach C suggests that temporal variations in rainfall help sustain egg populations during warmer months, particularly in July for the semi-arid region, October for the sub-tropical region, and April for the tropical region.

### Adult Mosquito Population Trends

Figure 3 shows variations in the population of infectious mosquitoes in different regions with differing transmission patterns across approaches. Approach A projects persistent infectious mosquito populations and Approach B suggests seasonal peaks in these populations, whereas shorter periods with abundance of infectious mosquitoes are observed in Approach C. Approach B suggests that while warmer temperatures in the tropical region increase mosquito mortality, they ultimately increase the risk of infection compared to the sub-tropical region, where populations drop substantially in winter but persist at high levles in the semi-arid and tropical regions. Approach C, however, shows that lower mosquito mortality rates in the sub-tropical region result in a sustained mosquito population, whereas warmer conditions in the semi-arid and tropical regions typically lead to higher mortality rates and as a result lower infectious mosquito populations. This approach further indicates that cooler temperatures in the sub-tropical region limit mosquito biting, while moderate temperatures in the tropical region increase the risk of mosquito infection. These populations persist throughout the year contributing to malaria transmission in these areas, particularly in semi-arid and tropical regions where these populations fluctuate rapidly. In the semi-arid and tropical regions, this approach produced short-lived transitory peaks in transmission, while stable seasonal dynamics were observed in the sub-tropical region where infectious mosquito populations persist for longer periods. Approach C further suggests that adult mosquito populations experience sharp, short-lived spikes throughout the years, which are characterized by irregular surges in infectious mosquito populations that remain very low across all regions during this period. These populations peak below 500 thousand mosquitoes in semi-arid and tropical regions, whereas infectious mosquito populations remain stable in the sub-tropical region. Increasing vector control coverage had the most pronounced effect under Approach A, with Approach B showing a slightly lesser impact. The overall transmission season remained remarkably stable despite higher coverage levels, especially in the sub-tropical region, where substantial reductions in infectious mosquito prevalence were observed. On the other hand, improved vector control coverage primarily influenced the intensity of peak mosquito infections, rather than altering the broader seasonal dynamics in Approach C.

**Fig. 3:**
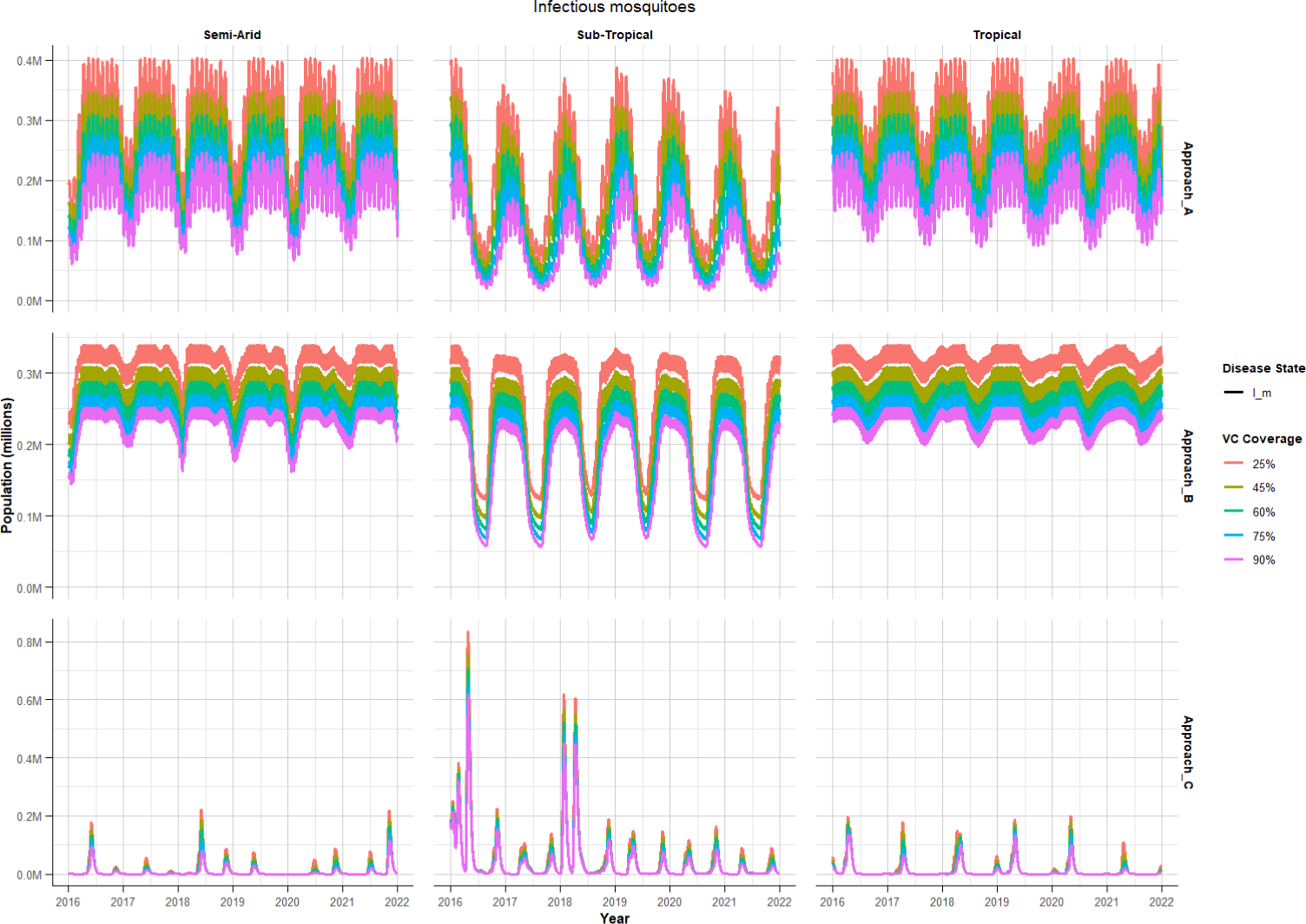
Projected infectious adult mosquito populations at different vector control coverage levles across regions.

### Host Population Dynamics

We observe in Figure 4 that all three modelling approaches project distinct patterns in the prevalence of asymptomatic and clinical malaria infections across climatic regions. In particular, Approach A consistently predicts a higher prevalence of asymptomatic infections relative to clinical cases which remain relatively low, peaking below 50 thousands across all regions. Approach B projected a similarly higher burden of asymptomatic infections, particularly in the semi-arid and tropical regions where transmission persisted through the winter months, despite substantial seasonal declines in the sub-tropical region. Approach C indicates short lived, multiple surges in asymptomatic and clinical cases throughout the year but no transmission occurs during the winter period. Asymptomatic populations in all regions stabilise around 300 thousand despite differing transmission seasons across Approaches. Approach A indicates that warmer temperatures in the semi-arid and tropical regions promote increased mosquito biting, which increases the risk of infection compared to the cooler sub-tropical region. However, Approach B indicates that lower mortality rates in the sub-tropical region enhance mosquito biting despite being restricted in the semi-arid and tropical regions due to higher mosquito mortality at warmer temperatures. Approach C suggests that adequate rainfall may sustain mosquito populations which may continue to transmit malaria infection even in regions with less favourable temperatures. Approach A projects annual peaks in asymptomatic populations which persist for longer in semiarid and tropical regions compared to sub-tropical region. In contrast, Approach B suggests that these populations are sustained for longer in the sub-tropical rather than the semi-arid tropical regions. Approach C predicts several irregular peaks in infectious mosquito populations throughout the year across all regions, which are driven by more frequent periods of optimal weather conditions as a result of ideal rainfall and temperature aligning to support mosquito development. The sporadic surges in infectious mosquito populations when Approach C is adopted, lead to sharp, short-lived peaks in new malaria cases in humans. Nonetheless, Approach B indicates that seasonal growth rather than irregular peaks in infectious mosquitoes, results in greater exposure and higher peaks in new malaria cases even compared to the persistent transmission in Approach A. Increasing vector control coverage produced the greatest impact under Approach A, where the prevalence of asymptomatic infections declined substantially, particularly within the sub-tropical region. Although Approach B also showed some reduction, these changes had minimal influence on the overall transmission season, which remained stable and relatively robust to higher coverage levels. In contrast, Approach C showed that increasing coverage primarily reduced the magnitude of peak asymptomatic infections.

**Fig. 4:**
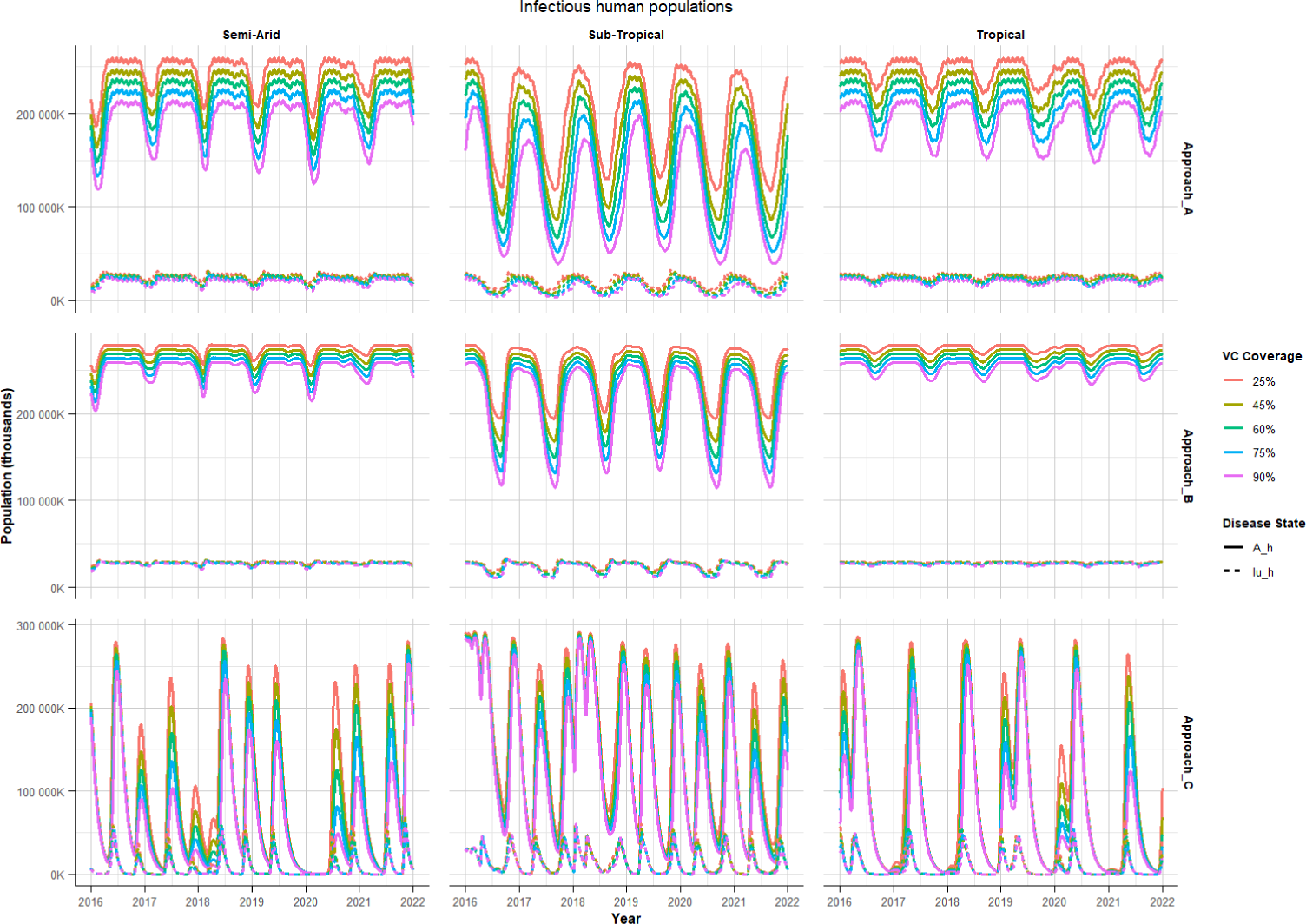
Projected infectious host populations at different vector control coverage levles across regions.

### Daily Malaria Incidence Projections

Figure 5 shows that all three approaches project different transmission seasons where Approach A projects sustained seasonal transmission peaking around 20 new cases per 1000 in the tropical region and 25 new cases per 1000 in semi-arid and sub-tropical regions. Improved vector control intervention from 25 % to 90 % annual coverage leads to reductions in new malaria cases, particularly in the sub-tropical region where a shift in the timing of peak transmission is observed. Approach B similarly projects new malaria cases peaking around 20 cases per 1000 in the tropical region and 25 cases per 1000 in the sub-tropical and semi-arid regions where annual preseason outbreaks are also observed. These peaks are more evident in the sub-tropical region where a significant reductions are observed in transmission during winter as a result of low temperatures, which limit contact between mosquitoes and humans in populations at risk which reduces the number of successful infectious bites during this period. Approach B also projects that malaria transmission persists during winter in the tropical region and semi-arid regions, as temperatures do not drop low enough to stop mosquito biting and as a result the spread of malaria infection. Approach C projects short lived peaks in new malaria cases and that transmission stops during winter across all regions, however this approach projects multiple peaks throughout the year. Irregular transmission is observed across regions, with cases peaking around 60 to 75 new cases per 1000 in tropical and semi-arid regions, whereas in the sub-tropical region where cases peak below 75 cases per 1000 and cases may rise up to 100 new cases per 1000 as experienced in 2018.

**Fig. 5:**
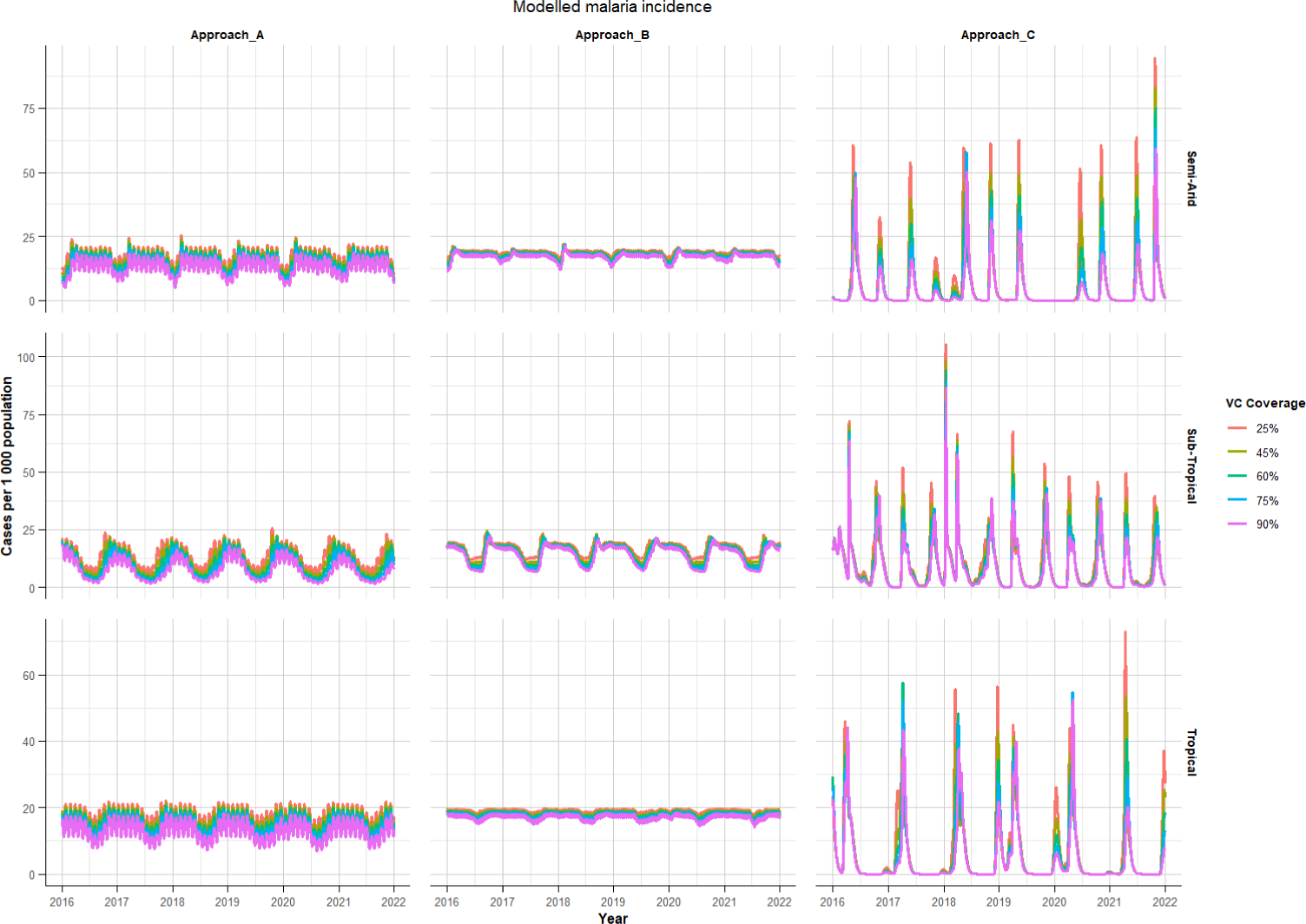
Projected daily malaria incidence rates (per 1000) at different vector control coverage levles across regions..

### Annual Malaria Transmission Trends

Figure 6 shows plots of annual trends in malaria transmission in semi-arid, sub-tropical and tropical regions during the 1950 to 2022 period. Approach A suggests that malaria incidence ranges between 40 to 60 new cases per 100 000 per year in semi-arid and tropical regions, with a slight increase throughout the simulation period. Whereas cases fluctuates between 20 to 50 new cases per 100 000 per year in the sub-tropical region despite reductions in annual cases as vector control coverage improves. Approach B suggests that annual cases in the sub-tropical region fluctuates fairly consistently below 60 cases per 100 000 and annual cases remain above 60 cases per 100 000 per year in the tropical and semi-arid regions. Approach C projects the least incidence with relatively stable transmission leading to a gradual rise in new cases peaking below 40 cases per 100 000 in all regions. Cases remain around 20 new cases per 100 000 per year in semi-arid and tropical regions, while cases may rise up to 40 new cases per 100 000 per year in the sub-tropical region. Although malaria transmission was halted around 1960 and again around 1980 in the sub-tropical region when vector control is sustained at 90 % annual coverage, malaria transmission persisted until 2022.

**Fig. 6:**
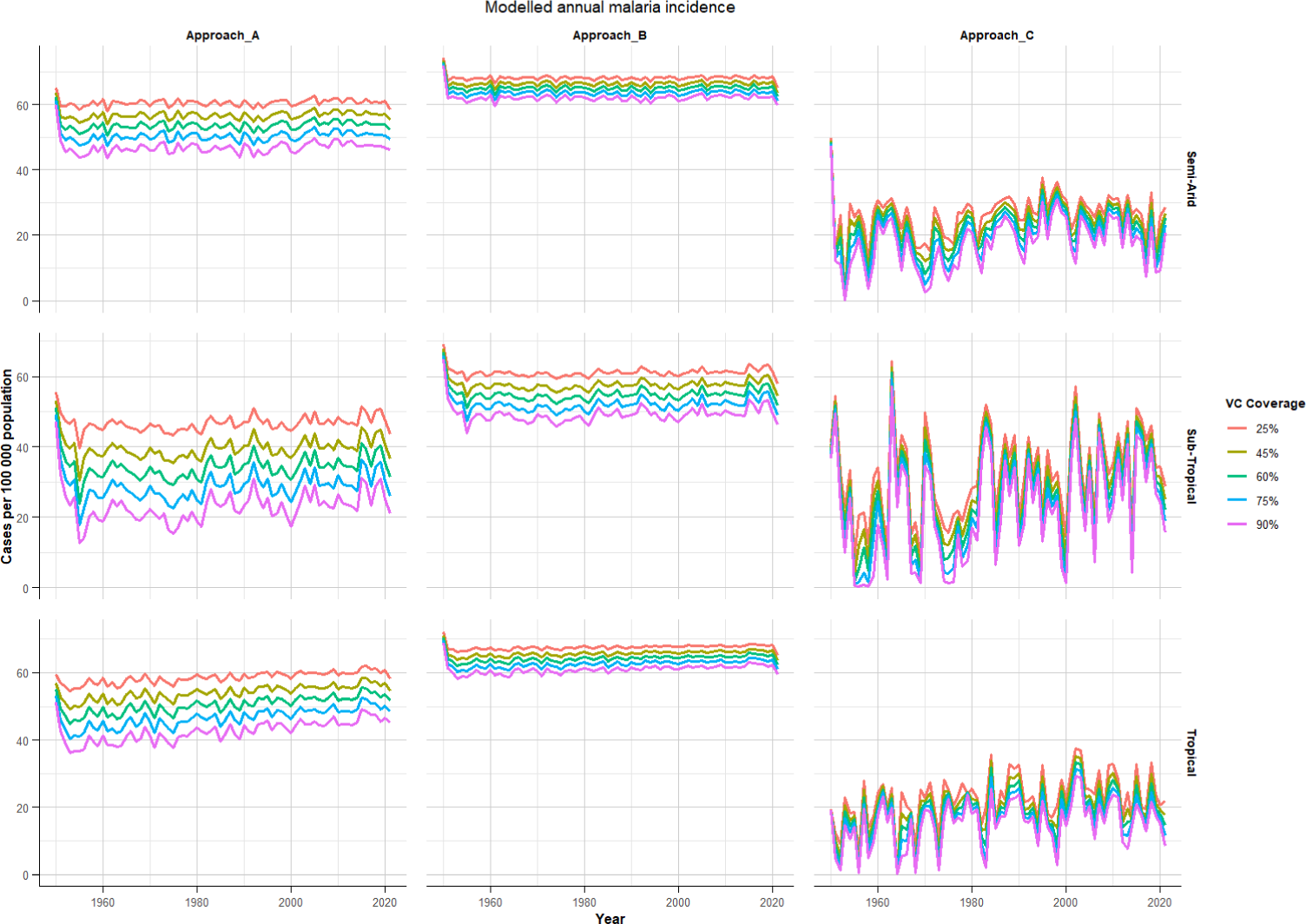
Projected annual malaria incidence rates (per 100 000) at different vector control coverage levles across regions.

## Discussion

This study used monthly mean rainfall and temperature datasets obtained from the Climate Change Knowledge Portal (CCKP) and were linearly interpolated to daily values for the 1950 to 2022 period in three areas representing semi-arid, tropical and sub-tropical regions. Model simulations were conducted and annual outcomes of malaria incidence are observed at this scale, however the seasonal patterns are explored at a shorter period from 2016 to 2022. A summary of the comparative analysis is provided in Table 3. Seasonal increases in asymptomatic populations play a key role in sustaining malaria transmission, particularly in Approach C, which demonstrates irregular peaks in adult mosquito populations. This pattern is influenced by temperature and rainfall, as higher temperatures accelerate mosquito development and parasite maturation, leading to an increased risk of malaria transmission. As observed in Approach A, moderate temperatures in the tropical region regulate mosquito survival, creating a larger mosquito population at risk compared to the sub-tropical region, which promotes more frequent biting in comparison to the semi-arid region. However, in the semi-arid region, as shown in Approach B, warmer temperatures accelerate mosquito biting, but the impact is less pronounced than in the tropical region. Approach C suggests that while rainfall plays a critical role in mosquito population dynamics, heavy rainfall periods in the semi-arid and tropical regions can inhibit mosquito growth by flushing out mosquito larvae, reduce mosquito populations and potentially limit the spread of malaria. On the other hand, insufficient rainfall reduces available breeding sites, which further lowers mosquito abundance and, consequently, malaria transmission. However, although Approach C projects sharp peaks in new daily cases, these are more sporadic and irregular. This pattern reflects the impact of rainfall on mosquito populations, as highlighted in previous studies: periods of heavy rainfall can flush out mosquito larvae, while insufficient rainfall limits breeding sites, both leading to fluctuating mosquito populations. Therefore, while Approach B’s seasonal growth in malaria cases indicates a more stable transmission cycle, the sporadic surges predicted by Approach C highlight the role of irregular environmental factors (e.g, heatwaves, flooding) in driving short-term increases in malaria incidence.

**Table 3:** Comparison of modelling approaches for malaria transmission dynamics.

| Aspect | Approach A | Approach B | Approach C |
| --- | --- | --- | --- |
| **Weather-Dependent Parameters** | Temperature-dependent biting rate, larval and adult mosquito mortality rate | Temperature-dependent parameters for fecundity, development, mortality, and biting rates | Rainfall-dependent environmental carrying capacity and aquatic survival; temperature-dependent development, mortality, and biting |
| **Regional Predictions** | Stable aquatic and adult populations supported year-round host infections, with limited mosquito biting and mortality contributing to the lowest incidence observed in the sub-tropical region | Large egg surges and peak adult populations generated seasonal host outbreaks, with higher annual cases in the semi-arid and tropical regions where extended warm periods sustaining transmission | Low but episodic egg populations produced sharp, sporadic peaks in daily incidence, driven by rainfall variability that influences breeding conditions and larval survival |
| **Environmental Drivers** | Temperature regulates mosquito survival and development, with moderate conditions supporting optimal growth and driving consistent transmission | Extended warm periods enhance mosquito development, survival, and biting activity, sustaining higher transmission potential | Irregular rainfall drives fluctuations, as heavy rains flush larvae while reduced rainfall restricts available breeding sites |
| **Transmission Patterns** | Endemic persistence with relatively stable adult mosquito populations, resulting in moderate fluctuations in incidence | Seasonal epidemics characterised by pre-season outbreaks, contributing to a higher annual burden in the sub-tropical region | Episodic outbreaks with sporadic surges closely linked to rainfall variability |
| **Vector Control Impacts** | High coverage reduces prevalence and incidence, but transmission persists, particularly in the tropical region | Control is effective in diminishing outbreak peaks, though it has limited impact on within-season fluctuations; this moderates seasonal peaks and reduces annual cases | Vector control substantially lowers incidence, but surges may persist due to asymptomatic reservoirs and environmental variability; at 90% coverage, multiple elimination episodes are observed |
| **Applicability to Real-World Scenarios** | Reflects endemic settings with stable transmission and persistent reservoirs, most applicable to regions with moderate climatic conditions | Represents seasonal, epidemic-prone regions influenced by extended warm periods | Captures the complexity of unstable transmission zones, where outbreaks are driven by rainfall variability and elimination is possible under sustained interventions |

Approach A projects the lowest daily incidence rates in the sub-tropical region due to cooler temperatures that limit mosquito biting, a finding consistent with past studies highlighting the role of temperature in regulating mosquito survival and development. Cooler temperatures reduce both mosquito and parasite maturation, limiting transmission, as supported by research indicating that mosquito development and malaria parasite maturation are hindered outside optimal temperature ranges [9, 12, 13, 15]. On the other hand, Approach B projects higher annual malaria cases in the semi-arid and tropical regions due to extended peak transmission periods. This pattern aligns with findings from past studies that associate warmer temperatures with accelerated mosquito development, which in turn prolongs the periods of high mosquito activity and malaria transmission [2, 12, 14, 15, 22]. This explains the higher cumulative cases under Approach B, where extended warm periods foster more consistent transmission. Approach C projects sharp peaks in new daily cases that are more sporadic and irregular, reflecting fluctuating mosquito populations driven by rainfall variability. Heavy rainfall can wash away mosquito larvae, reducing mosquito numbers, while insufficient rainfall results in fewer breeding sites, both contributing to the irregular surges seen in Approach C. This pattern highlights the importance of process-based models that consider both temperature and rainfall impacts on mosquito and parasite development. Dynamic models of malaria transmission, which account for these interactions provide key insights into the duration of transmission periods as well as transmission intensity and frequency. These explain how prolonged warm conditions can sustain malaria transmission over time, as observed in Approach A and B, and how environmental fluctuations contribute to the sporadic surges in Approach C. Although rainfall has often been considered a secondary factor in malaria transmission, these results highlight its critical role in shaping mosquito populations, especially in regions with irregular rainfall patterns, as seen in Approach C.

Effective vector control targeting adult mosquitoes reduces annual incidence rates across all regions, with Approach A showing the most substantial reduction. A mechanistic model calibrated with weather data from various African regions found that malaria infection rates rise when temperatures range from 16 to 25_0_C but decrease when temperatures exceed 25_0_C. In the semi-arid region, temperatures peaked around 30_0_C, creating a challenging environment for mosquito survival. This suggests that adult mosquito populations, which are crucial for transmission, are most abundant and active within the lower range of these temperatures, supporting the substantial reductions in transmission observed under Approach A. Studies have shown that temperatures above 28_0_C tend to reduce transmission due to the stress on both mosquito populations and the malaria parasite [20, 27, 31]. However, the combination of high temperatures and intermittent rainfall results in a variable environment, allowing mosquito populations to thrive during wetter periods, leading to seasonal peaks in transmission, as seen in Approach C. These irregular surges in transmission reflect the fluctuating nature of the environment in this region.In contrast, the tropical region, with temperatures ranging from 25_0_C to 30_0_C and the highest rainfall (250 to 300 mm), creates optimal conditions for mosquito breeding. The malaria parasite also matures best at temperatures between 21 and 25_0_C, suggesting that the tropical region likely experiences a consistent transmission cycle during heavy rainfall months. This consistency is reflected in Approach B, where seasonal variability in annual cases corresponds to favourable environmental conditions. Approach B also shows a notable reduction in incidence, although seasonal variability influences annual cases. The high rainfall and suitable temperatures provide ideal conditions for both the aquatic stages of the mosquito life cycle and the adult mosquito populations, resulting in higher malaria incidence in this region. This is consistent with findings from climate-based vector-host models reveal that transmission is optimised when temperatures are between 21 and 25_0_C, and rainfall ranges from 95 to 125 mm, conditions that support mosquito breeding [2, 6, 7, 21, 22]. Seasonal transmission patterns observed in Approach B likely reflect these environmental factors, with transmission peaking during favourable periods and waning during less optimal times. These findings emphasise the importance of sustained vector control in regions where malaria persists during cooler periods to prevent resurgence when conditions become favourable.

Increasing vector control coverage to 90% results in periods of low to no transmission, particularly in the tropical region when Approach C is adopted. This aligns with the results from vector-host models, which indicate that high levels of control can significantly reduce mosquito populations, leading to low transmission. However, as seen in Approach C, resurgences in malaria cases occur after periods of low transmission, driven by sustained asymptomatic populations and irregular spikes in infectious mosquitoes. These resurgences reflect findings from studies that show the seasonal fluctuations in larval density and mosquito breeding success, with cooler temperatures hindering mosquito activity, particularly biting, and slow parasite development [7, 12, 26, 31, 32]. The sub-tropical region, with temperatures around 15_0_C, is less conducive to year-round mosquito survival and transmission. Additionally, with the least rainfall, peaking below 150 mm, this region has fewer breeding sites, further contributing to lower transmission rates. However, despite these lower environmental risks, malaria transmission can still occur, particularly with asymptomatic individuals or seasonal surges in mosquito populations, which is reflected in the sporadic surges predicted in Approach C. All three approaches indicate low malaria infection rates within mosquito populations across all regions, suggesting that vector control measures are effective at reducing the parasite load in mosquitoes. However, Approach A projects a consistent adult mosquito population throughout the year, while Approach B suggests a seasonal pattern with localized peaks, and Approach C shows sporadic, localized surges driven by mismatches in temperature and rainfall seasonal patterns. Despite these differences, all approaches indicate large, sustained populations at risk of infection, emphasizing the ongoing need for comprehensive vector control strategies that account for environmental factors to address both aquatic and adult mosquito populations. These modelling approaches have shown that changes in environmental factors like temperature and rainfall can shift the suitability of an area for malaria transmission, sometimes causing outbreaks in previously unaffected regions. The seasonal growth in malaria cases under Approach B contrasts with the sporadic surges under Approach C, where the influence of rainfall and environmental fluctuations is more pronounced, illustrating the complex and dynamic nature of malaria transmission in different regions.

Although this study provides important insights into how weather conditions influence malaria transmission, it has several limitations. The modelling approaches account for specific assumptions regarding the relationship between mosquito life cycles and environmental factors. Additionally, relying on monthly mean weather data without accounting for daily fluctuations may restrict the applicability of the model to localized regions and limit its utility for short-term forecasting of transmission periods. Moreover, the models primarily emphasize weather influences while neglecting critical factors such as vector and parasite species diversity, healthcare access, and human movement, which significantly impact malaria dynamics. A key limitation is the use of parameter formulations derived from multiple vector species, which may reduce the accuracy of the model in capturing the seasonal dynamics specific to any single species. Future research should aim to develop more comprehensive malaria control strategies by examining the effectiveness of treatments, larval source management, and the impact of case importation due to travel. Incorporating these elements into the models and validating them with vector surveillance data on abundance and distribution would enhance their practical relevance for malaria prevention and control.

## Conclusions

This comprehensive analysis underscores the importance of context-specific malaria control strategies and enhances our understanding of how temperature and rainfall influence various factors in malaria transmission dynamics, especially in regions with variable weather conditions. The three modelling approaches focus on distinct aspects of malaria transmission, highlighting the importance of tailoring interventions to specific local conditions, such as rainfall patterns, vector behaviour, and the impacts of climate change. The simulation results indicate that climatic fluctuations such as heat shocks or flooding can trigger surges in mosquito populations and subsequent malaria outbreaks, even with high levels of vector control. This has potentially far-reaching consequences because extreme weather events may enable the introduction of malaria into malaria-free areas or areas recently cleared of the disease. However, the findings also show that consistent and effective vector control significantly reduces malaria incidence, particularly in regions with moderate temperatures. Effective malaria control requires the integration of weather predictions into intervention plans to maintain efficacy in the face of changing environmental conditions. To accurately predict malaria transmission dynamics and inform intervention strategies, malaria control programs should prioritize selecting climate-vector modelling frameworks that are tailored to local climatic and transmission conditions. In light of these considerations, the following recommendations are proposed to address challenges and ensure sustained malaria control amidst evolving weather patterns:

1. Malaria control programs should select modelling structures (i.e., Approaches A, B, and C) that reflect the local interplay between temperature, rainfall, and vector ecology.
2. Supplementary interventions, such as larval source management, should be incorporated to enhance the effectiveness of primary vector control measures.
3. Accurately modelling seasonal variations in mosquito abundance, infection risk, and peak transmission periods is critical for optimizing the timing, targeting, and scale of vector control interventions, particularly to achieve and maintain coverage levels above 90% annually.
4. In settings with overlapping climatic drivers or ecological uncertainty, fitting modelling approaches to local data can provide a more accurate reflection of transmission risk and intervention outcomes.
5. Future modelling studies should integrate critical factors such as human mobility, healthcare access, and species diversity in both vectors and parasites to understanding localized outbreaks and long-term elimination prospects.

By selecting the appropriate climate-vector model, malaria control programs can ensure effective and targeted interventions, mitigating the impacts of environmental fluctuations on malaria transmission dynamics.

## Supporting information

AppendixII

AppendixI

## List of Abbreviations

WHO: World Health Organization
ACTs: Artemisinin-based Combination Therapies
IRS: Insecticide Residual Spraying
LLINs: Long-Lasting Insecticide Nets
ODEs: Ordinary Differential Equations
CCKP: Climate Change Knowledge Portal
NLS: Nonlinear Least Squares

## Declarations

### Ethics Approval

The research protocol for this study was reviewed and approved by the University of Cape Town’s Faculty of Health Sciences Human Research Ethics Committee (UCT HREC) **[058/2024]** and the Limpopo Provincial Department of Health **[LP 2024-05-026]**.

### Consent for publication

Not applicable.

### Data Availability

The temperature and rainfall datasets used in the current study are publicly available in the World Bank, Climate Change Knowledge Portal repository. https://climateknowledgeportal.worldbank.org

### Code Availability

The code developed to generate simulation results for the model is publicly available at the Malaria Weather Models Github repository. https://github.com/maditogladstone/regionalmalariaweathermodels.git

### Author Contributions

GT developed the mathematical model, implemented the code, conducted the analysis, and prepared the manuscript draft. SP contributed to the conceptual design of the study, provided guidance throughout the research process. SP also reviewed and provided feedback on the manuscript. BD reviewed, revised and provided feedback on the manuscript. All authors read and approved the final manuscript.

## Acknowledgements

This research was financially supported by the Bill and Melinda Gates Foundation (BMGF). We express our gratitude for the insights and resources provided by the Malaria Modelling and Analytics, Leaders in Africa (MMALA) programme. Additionally, we extend our thanks to the Limpopo Malaria Control Programme, Clinton Health Access Initiative (CHAI) South Africa and the SADC Elimination 8 (E8) Initiative Technical Working Group (TWG) for their invaluable support and collaboration throughout this study.

