## AppendixII for "Comparing Three Approaches to Modelling the Effects of Temperature and Rainfall on Malaria Incidence in Different Climatic Regions"

### Appendix II: Supplemental Plots

#### Transmission Parameter Curves by Approach

The following functions represent how temperature affects key transmission parameters. For Approach A, the mosquito biting rate ($a$) is derived from studies on *Anopheles pseudopunctipennis* vectors [1, 26, 43, 44], and the mortality rates ($\mu_l$ and $\mu_m$) was defined using data from *Culex quinquefasciatus and Aedes aegypti* mosquitoes [45]. Figure 2 provides plots showing how these parameters change with temperature.

$$\mu_l = \max\left(0, \frac{1}{-4.4 + 1.31(T+2) - 0.03(T+2)^2}\right)$$

$$\mu_m = \frac{3.04}{30.4} + \frac{29.564}{30.4} \times \exp\left(-\frac{T + 273.15 - 278}{2.7035}\right)$$

$$a = \frac{1.25}{107.204 - 13.3523T + 0.677509T^2 - 0.0159732T^3 + 0.000144876T^4}$$

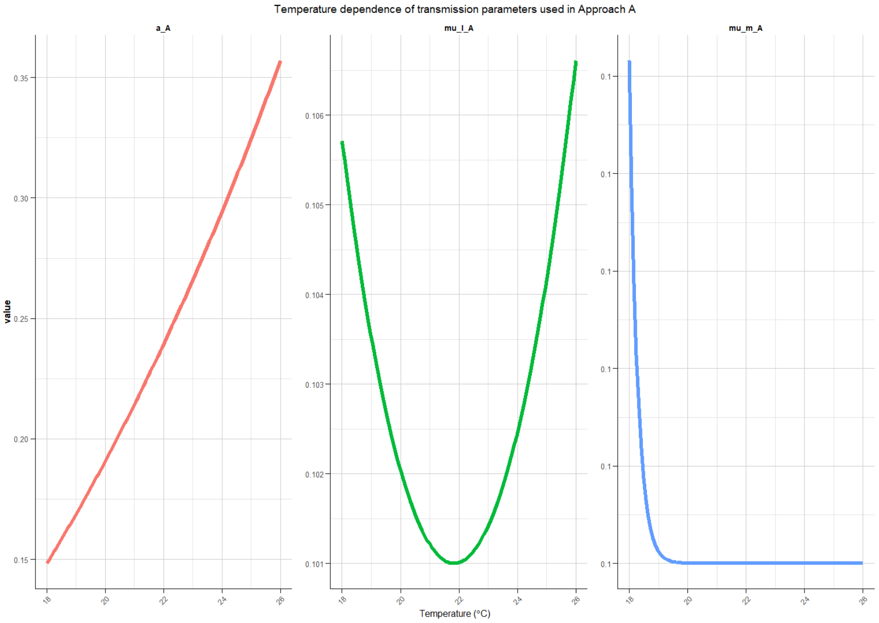


**Fig. 2**: Model parameters used for Approach A.

Figure 3 shows thermally driven changes in egg production ($n_e$), oviposition ($\theta$), development ($\kappa_e$, $\kappa_l$, and $\kappa_p$), and mortality ($\mu_e$ and $\mu_l$), were parameterized for $A$.

*arabiensis* [22, 39] in Approach B. These processes were captured through cubic polynomial functions constrained to biologically plausible values. The mosquito biting rate ($a$) follows a non-linear thermal response drawing on empirical relationships for *Anopheles spp.* and *A. pseudopunctipennis* [22, 41]. These parameters are represented by the following functions.

$$
\begin{aligned}
n_e &= -3.387987 + 1.038248 \times \left(-0.61411T^3 + 38.93T^2 - 801.27T + 5391.4\right) \\
\theta &= -7.648455 + 1.147739 \times \left(0.00054T^3 - 0.038T^2 + 0.88T\right) \\
\kappa_e &= -0.3485345 - 0.2186501 \times \left(0.012(T+2)^3 - 0.81(T+2)^2 + 18(T+2) - 135.93\right) \\
\mu_e &= 0.2874394 - 0.1396794 \times \left(0.0033(T+2)^3 - 0.23(T+2)^2 + 5.3(T+2) - 40\right) \\
\kappa_l &= 0.4722003 - 0.09495794 \times \left(-0.002(T+2)^3 + 0.14(T+2)^2 - 3(T+2) + 22\right) \\
\mu_l &= 1.115093 - 0.1583374 \times \left(0.00081(T+2)^3 - 0.056(T+2)^2 + 1.3(T+2) - 8.6\right) \\
\kappa_p &= 0.3881068 + 0.2755463 \times \left(-0.0018(T+2)^3 + 0.12(T+2)^2 - 2.7(T+2) + 20\right) a \quad = 2.5 \times 0.000203 \times (
\end{aligned}
$$

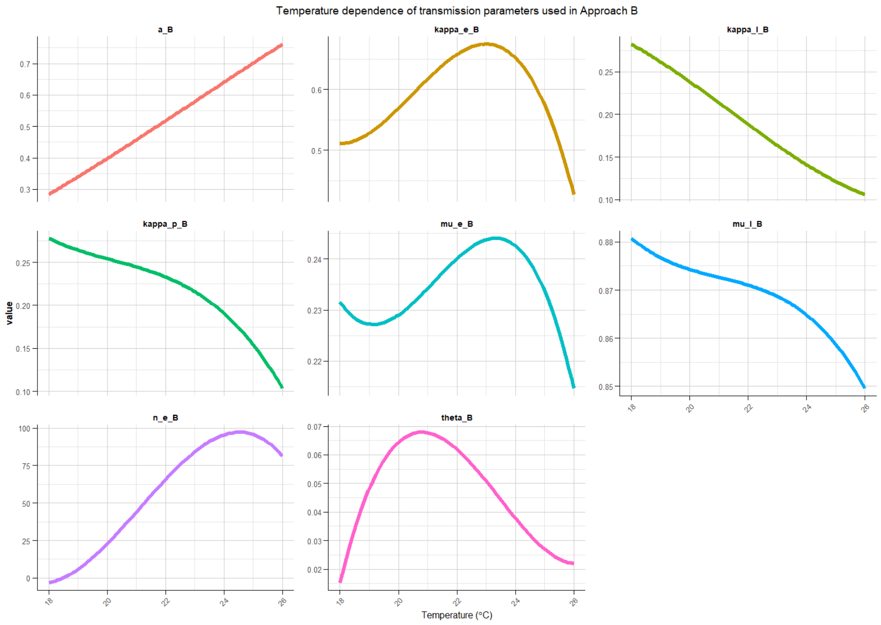


**Fig. 3**: Model parameters used for Approach B.

Figure 4 shows how in Approach C, the larval mortality rate ($\mu_l$), adult mortality rate ($\mu_m$), parasite development rate ($\gamma_m$), and biting rate ($a$) are defined using the following non-linear thermal functions. These functions were parameterized from studies of *Anopheles spp.* and the *A. gambiae* complex [22, 25, 40, 42], alongside biting rate relationships informed by *A. pseudopunctipennis* [22, 41].

$$\mu_l = 0.0025(T+2)^2 - 0.094(T+2) + 1.0257$$
$$\mu_m = \max\left(0, \frac{1}{-4.4 + 1.31(T+2) - 0.03(T+2)^2}\right)$$
$$\gamma_m = \max\left(0, \frac{T-16}{111}\right)$$
$$a = 7.5 \times 0.000203 \times (T^2 - 11.7T) \times \sqrt{42.3 - T}$$

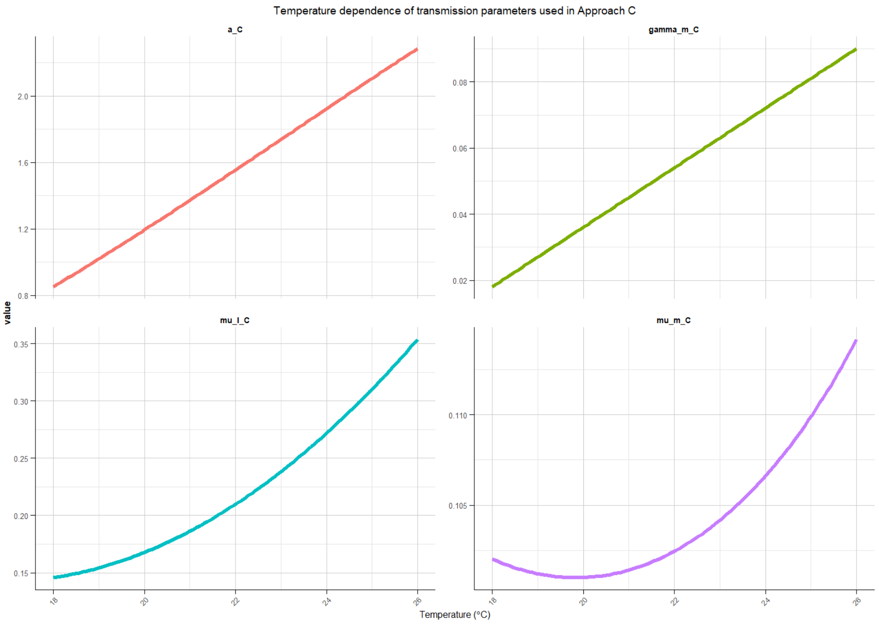


**Fig. 4**: Model parameters used for Approach C.

#### Surface Responses to Temperature and Rainfall (Approach C)

Figure 5 shows that the carrying capacity ($K_m$) for mosquito breeding increases linearly with rainfall, reflecting resource availability. The following function defines the carrying capacity as a function of rainfall.

$$K_m = \frac{100}{0.01} \cdot R = 10^4 \cdot R$$

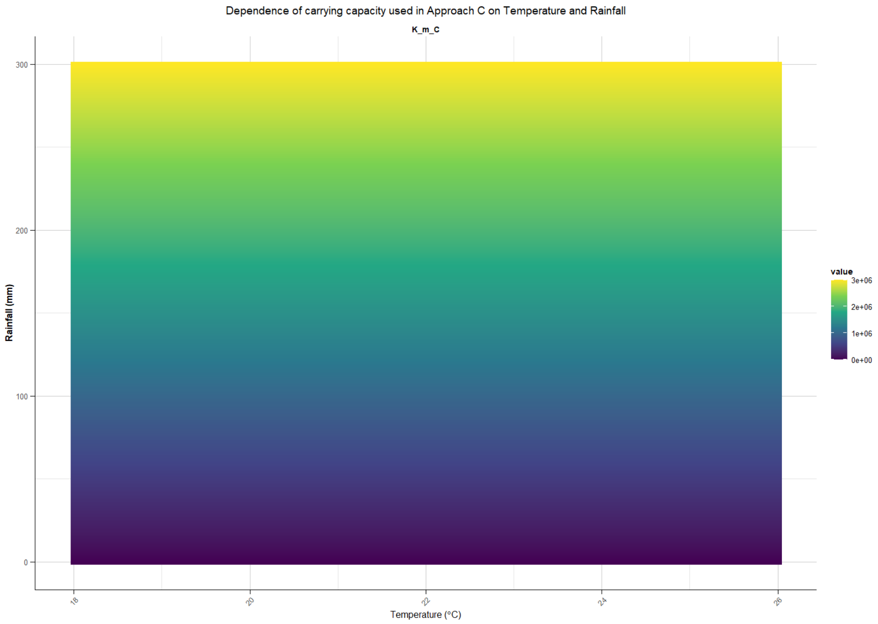


**Fig. 5**: Adult carrying capacity profile used for Approach C.

Surfaces of egg, larval, and pupal development rates ($\kappa_e$, $\kappa_l$, and $\kappa_p$) derived from studies on *A. gambiae* complex mosquitoes show complex interactions between climatic variables under Approach C [22, 25, 40, 42]. The developmental rates of aquatic stages as functions of temperature and rainfall given by the following functions.

$$\kappa_e = \min\left(1, \frac{4 \cdot \max p_e}{R_l^2} \cdot R \cdot \max(0, R_l - R)\right)$$
$$\kappa_l = \exp\left(-\frac{1}{0.0557(T+2) - 0.06737}\right) \cdot \min\left(1, \frac{4 \cdot \max p_l}{R_l^2} \cdot R \cdot \max(0, R_l - R)\right)$$
$$\kappa_p = \min\left(1, \frac{4 \cdot \max p_p}{R_l^2} \cdot R \cdot \max(0, R_l - R)\right)$$

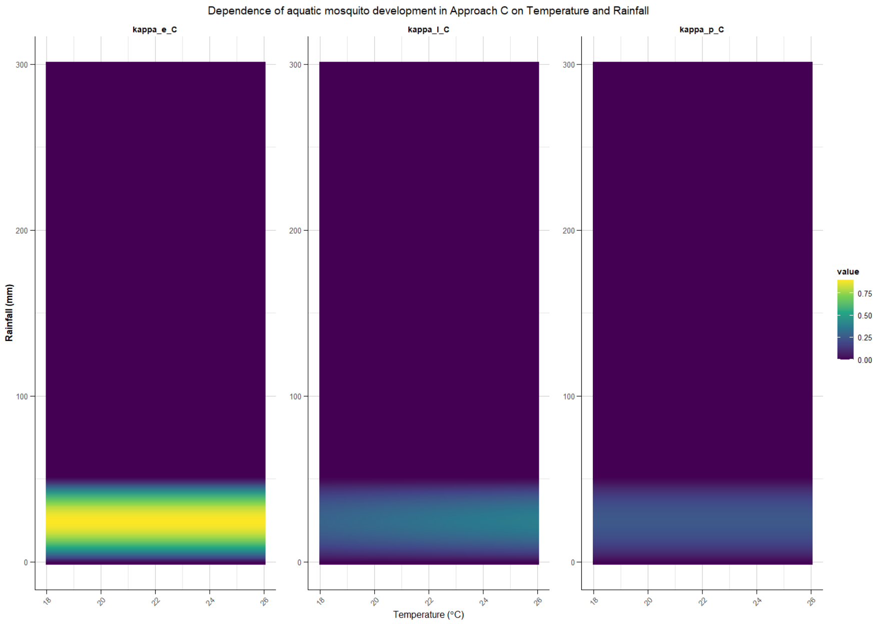


**Fig. 6**: Survival rate profiles for eggs, larvae and pupae used for Approach C.

#### Climate Data

Figure 7 provides a timeseries of mean monthly temperature trends from the Climate Change Knowledge Portal (CCKP) are shown across the semi-arid, tropical, and sub-tropical regions.

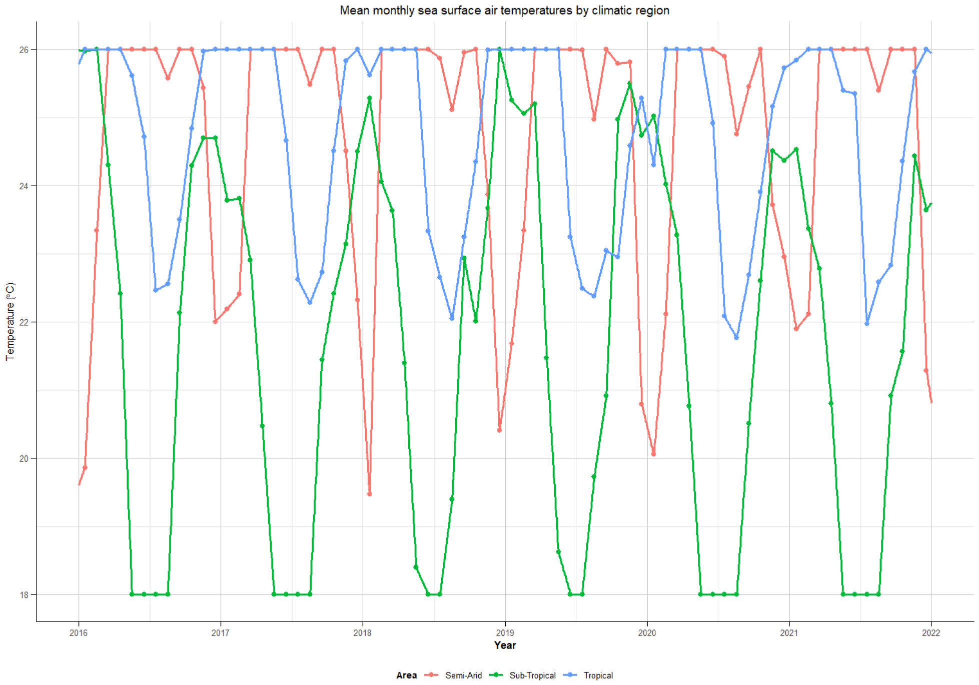


**Fig. 7**: Temperature trends for the 2016 to 2022 period

Figure 8 provides a timeseries of mean monthly rainfall trends from the Climate Change Knowledge Portal (CCKP) are shown across the semi-arid, tropical, and sub-tropical regions.

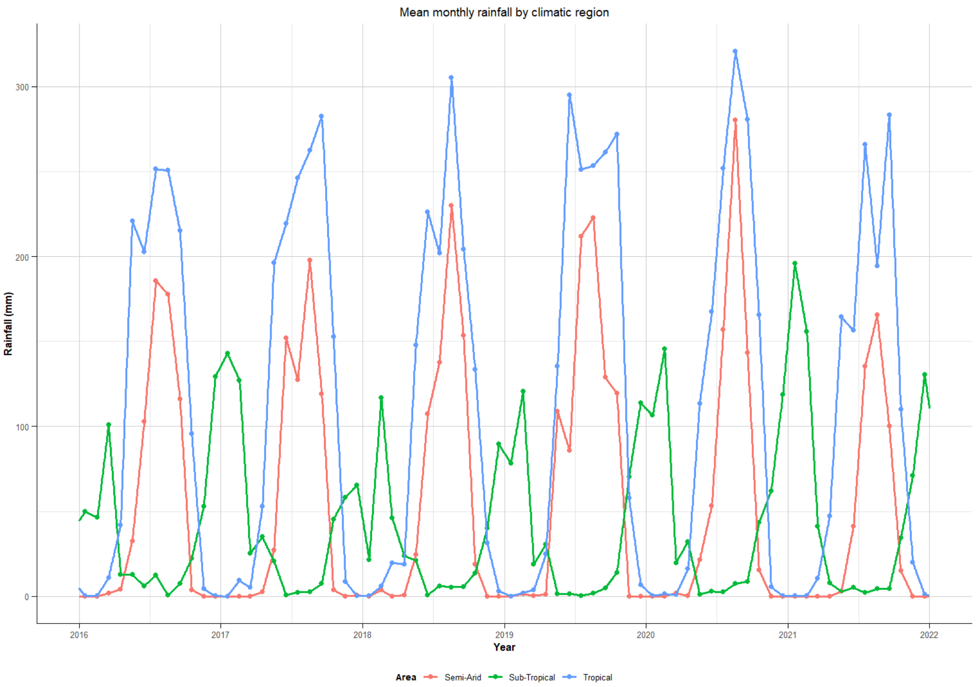


**Fig. 8**: Rainfall trends for the 2016 to 2022 period

#### Time-Series of Derived Parameters by Approach

Temperature-derived values of biting rate and mortality over time across semi-arid, tropical, and sub-tropical regions.

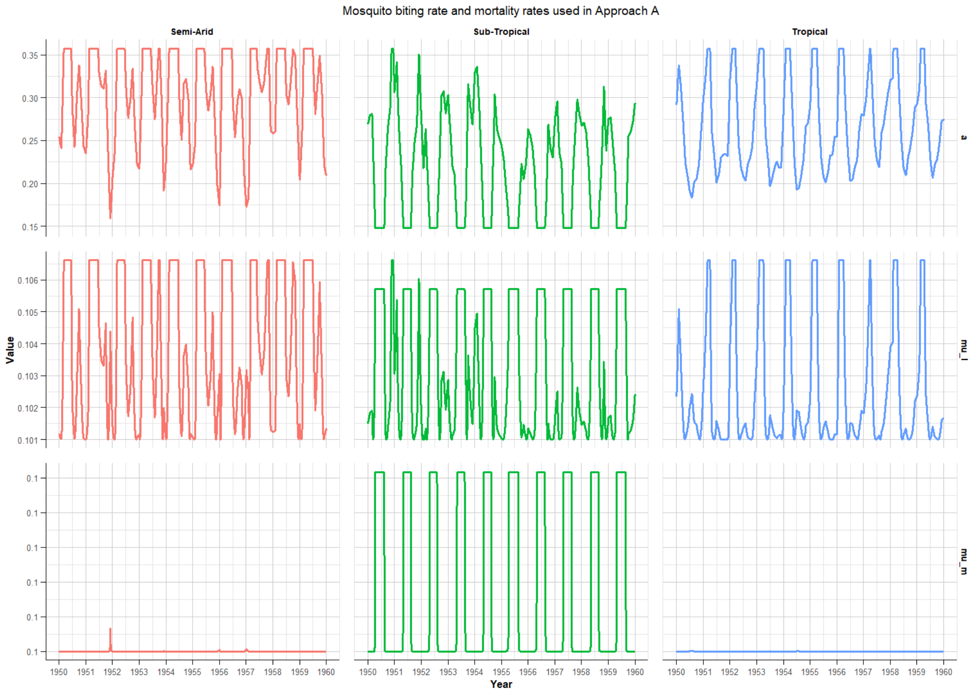


**Fig. 9**: Model parameters used for Approach A.

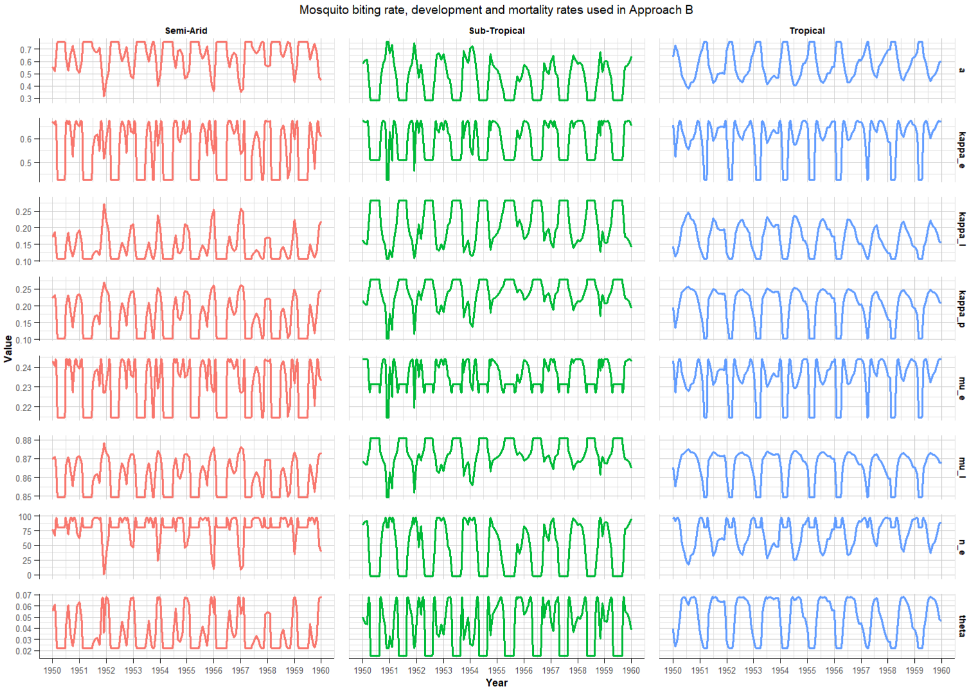


**Fig. 10**: Model parameters used for Approach B.

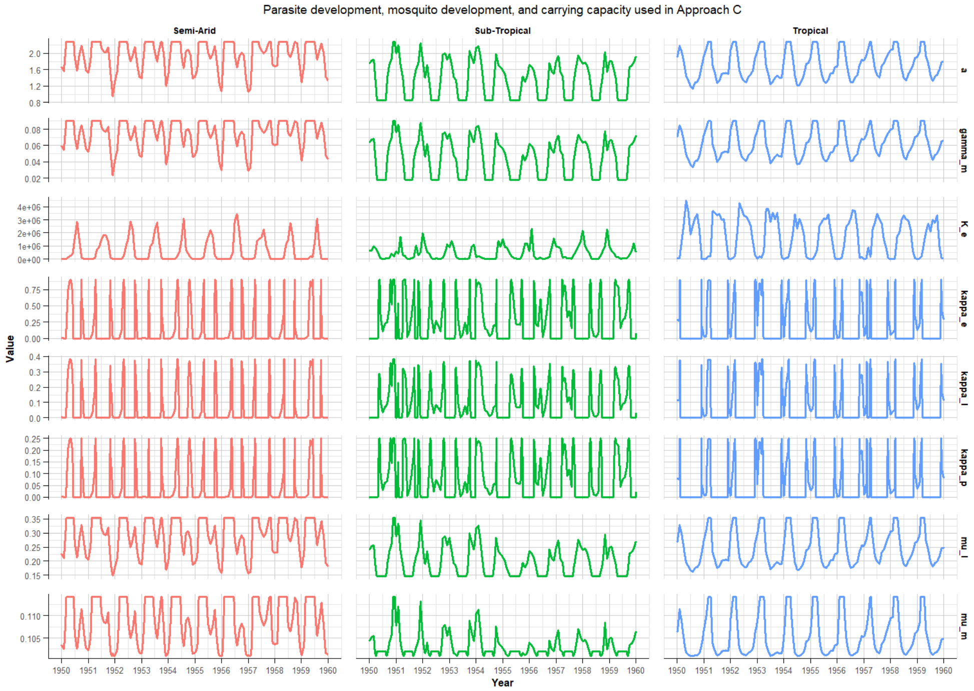


**Fig. 11**: Model parameters used for Approach C.
