## AppendixI for "Comparing Three Approaches to Modelling the Effects of Temperature and Rainfall on Malaria Incidence in Different Climatic Regions"

### Appendix I: Model Equations and Fitting Methods

#### Model Equations

The mathematical model developed in the study is represented by the following systems of equations.

**Aquatic mosquito population:**

$$\frac{dE_a(t)}{dt} = n_e\theta\left(1 - \frac{M(t)}{K_m}\right)M(t) - \kappa_e E_a(t) - \mu_e E_a(t) \tag{1}$$

$$\frac{dL_a(t)}{dt} = \kappa_e E(t) - \kappa_l L_a(t) - \mu_l L_a(t) \tag{2}$$

$$\frac{dP_a(t)}{dt} = \kappa_l L_a(t) - \kappa_p P_a(t) - \mu_p P_a(t) \tag{3}$$

**Adult mosquito population:**

$$\frac{dS_m(t)}{dt} = \kappa_p P_a(t) - \lambda_m S_m(t) - \mu_m S_m(t) \tag{4}$$

$$\frac{dE_m(t)}{dt} = \lambda_m S_m(t) - \gamma_m E_m(t) - \mu_m E_m(t) \tag{5}$$

$$\frac{dI_m(t)}{dt} = \gamma_m E_m(t) - \mu_m I_m(t) \tag{6}$$

**Human population:**

$$\frac{dS(t)}{dt} = \mu_h P(t) - \lambda_h S(t) + \rho R(t) - \mu_h S(t) \tag{7}$$

$$\frac{dE(t)}{dt} = \lambda_h S(t) - \gamma_h E(t) - \mu_h E(t) \tag{8}$$

$$\frac{dA(t)}{dt} = p_a\gamma_h E(t) + \omega I_u(t) - \delta_r A(t) - \mu_h A(t) \tag{9}$$

$$\frac{dI_u(t)}{dt} = \eta + (1 - p_a)\gamma_h E(t) - \omega I_u(t) - \nu I_u(t) - \delta_r I_u(t) - \tau_u I_u(t) - \mu_h I_u(t) \tag{10}$$

$$\frac{dI_s(t)}{dt} = \nu I_u(t) - \alpha I_s(t) - \tau_s I_s(t) - \mu_h I_s(t) \tag{11}$$

$$\frac{dT_u(t)}{dt} = \tau_u I_u(t) - \delta_u T_u(t) - \mu_h T_u(t) \tag{12}$$

$$\frac{dT_s(t)}{dt} = \tau_s I_s(t) - \delta_s T_s(t) - \mu_h T_s(t) \tag{13}$$

$$\frac{dR(t)}{dt} = \delta_r A(t) + \delta_r I_u(t) + \delta_u T_u(t) + \delta_s T_s(t) - \rho R(t) - \mu_h R(t) \tag{14}$$

**Intervention coverage:**

$$\frac{dV(t)}{dt} = \sigma - \zeta V(t) \tag{15}$$

where $\sigma$ represents the annual rate of vector control coverage and $\zeta$ determines the decay rate of intervention coverage. The forces of infection in the mosquito ($\lambda_m$) and human ($\lambda_h$) populations for the model are defined respectively by Equation 16 and 17.

$$\lambda_m = \frac{ab}{1+\xi V(t)} \times \frac{I_h(t)}{P(t)} \tag{16}$$

$$\lambda_h = \frac{ac}{1+\xi V(t)} \times \frac{M(t)}{P(t)} \times \frac{I_m(t)}{M(t)} \tag{17}$$

where $\xi$ determines the efficacy of vector control interventions.

**Table 1**: Parameter descriptions and baseline values (per day) used to simulate the model.

| Symbol | Description | Value | Source |
|---|---|---|---|
| $n_e$ | Average number of eggs laid | 12 | [2, 22, 25] |
| $\kappa_e$ | Egg hatching rate | 0.30 | [12, 22] |
| $\mu_e$ | Egg mortality rate | 0.10 | [12, 22] |
| $\kappa_l$ | Larval development rate | 0.06 | [12, 22] |
| $\mu_l$ | Larval mortality rate | 1.0 | [12, 22] |
| $\kappa_p$ | Pupal development rate | 0.70 | [12, 22] |
| $\mu_p$ | Pupal mortality rate | 0.40 | [12, 22] |
| $\mu_m$ | Mortality rate in adult mosquitoes | 0.104 | [37] |
| $\gamma_m$ | Mosquito incubation rate | 0.1 | [37] |
| $\mu_b$ | Birth rate of humans | 0.03 | [2, 22] |
| $\mu_h$ | Mortality rate in humans | 0.01 | [2, 22] |
| $\delta_u$ | Recovery rate for uncomplicated symptoms | 0.03 | [2, 25, 37] |
| $\delta_s$ | Recovery rate for severe symptoms | 0.02 | [2, 25, 37] |
| $\gamma_h$ | Incubation rate within humans | 0.08 | [2, 25, 37] |
| $\nu$ | Development rate of severe symptoms | 0.05 | [37] |
| $\tau_u$ | Treatment rate for uncomplicated symptoms | 0.1 | [12] |
| $\omega$ | Recovery rate from severe symptoms | 0.2 | [37] |
| $\tau_s$ | Treatment rate for severe symptoms | 0.15 | [12] |
| $\mu_s$ | Disease-induced mortality rate | 0.05 | [25, 37] |
| $\rho$ | Loss of temporary immunity | 0.1 | [37] |
| $\sigma$ | Annual vector control coverage | 0.25 - 0.90 | - |
| $\zeta$ | Vector control operational efficacy period | 365 | - |

#### Parameter Adjustment and Functional Fitting Procedure

The temperature-dependent relationships adopted from [22] were derived from fitted empirical curves intended to represent mosquito life-history processes. However, these functions may not fully capture biologically realistic dynamics across the entire temperature range. Specifically, the functional forms for egg development ($\kappa_e$), pupal development ($\kappa_p$), and pupal mortality ($\mu_p$) rates yield negative values under certain temperature conditions. In particular, $\mu_p$ remains strictly negative across the 18 to 26 °C temperature range, while $n_e$ predicted increased egg production at temperature below 18 °C, indicating that further adjustment or reparameterisation were necessary to ensure biological feasibility. To address this, each rate was re-estimated using nonlinear least squares (NLS) regression to ensure that temperature thresholds, inflection points, and asymptotic behavior are biologically meaningful and consistent across life stages. NLS allows flexible curve fitting to nonlinear biological processes, which are rarely well-described by linear models. It enables parameter estimation grounded in observed data, while preserving the functional form needed for mechanistic modeling, which helps correct implausible outputs (e.g., negative rates or unrealistic peaks) that may arise from extrapolating original fits beyond their intended range. Each life-stage rate (e.g., development, mortality, or egg-laying rate) was re-estimated using NLS regression model based on the general form: rate $\sim a + b \times f(T)$, where $a$ and $b$ are scaling coefficients estimated from the data, $f(T)$ represents the original temperature-dependent function from [22], and $T$ is temperature (°C). The model was fitted using the `nls()` function in R to obtain coefficients $a$ and $b$ that best matched the literature-based data. The results of the fitting process are presented in Figure 1. Predicted rates were generated over a temperature grid (18 to 26 °C) using the fitted model. Negative predictions were truncated to zero to prevent biologically implausible values, while parameters (except for the number of eggs laid, $n_e$) were capped at 1 to maintain dimensional consistency with probabilities or proportions (e.g., $\kappa_e$, $\mu_l$, or $\theta$). For egg-laying, an additional biological constraint was applied such that no egg production occurred below 18 °C, consistent with laboratory observations of temperature-dependent oviposition thresholds [25, 36, 39–42].

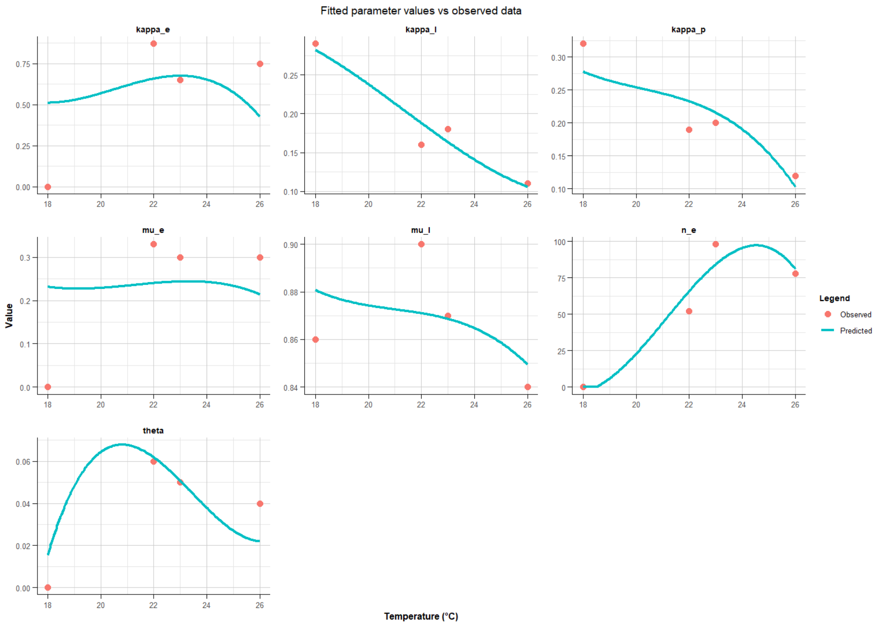

**Fig. 1**: Observed data vs adjusted model parameters used in Approach B.
